# Improving case detection through TB contact risk stratification by Xpert MTB/RIF Ultra and spatial parameters. Evaluation of an innovative ACF strategy in Mozambique

**DOI:** 10.1101/2022.10.18.22281174

**Authors:** Belén Saavedra, Dinis Nguenha, Laura de la Torre-Pérez, Edson Mambuque, Gustavo Tembe, Laura Oliveras, Matthew Rudd, Paulo Philimone, Benedita Jose, Juan Ignacio Garcia, Neide Gomes, Shilzia Munguane, Helio Chiconela, Milton Nhanommbe, Santiago Izco, Sozinho Acacio, Alberto L. García-Basteiro

## Abstract

Prompt diagnosis is critical for tuberculosis (TB) control, as it enables early treatment which in turn, reduces transmission and improves treatment outcomes. We aimed to determine the impact of the scale-up of Xpert Ultra as frontline test for TB diagnosis, and an innovative active-case finding (ACF) strategy (based on Xpert Ultra semi quantitative results and spatial parameters) on new TB cases diagnosed in a semi-rural district of Southern Mozambique. From January-December 2018 we recruited all incident TB-cases (index cases, ICs) and their household contacts (HCs). Community contacts (CCs) recruitment depended on the semi-quantitative results of Xpert Ultra of the IC and the population density of the area where the IC lived in. TB-contacts, either symptomatic or people living with HIV (regardless of symptoms), were asked for providing a spot sputum for lab-testing. Trends on TB case notification in the intervention area were compared to the previous years and to those of two districts in the south of the Maputo province (control area) using an interrupted time series analysis with and without control (CITS/ITS). A total of 1010 TB ICs (37.2% laboratory-confirmed) were recruited; 3165 HC and 4730 CC were screened for TB. Eighty-nine additional TB cases were identified through the ACF intervention (52.8% laboratory-confirmed). The ACF intervention increased by 8.2% all forms of TB cases detected in 2018. CITS model showed an increase of laboratory confirmed TB cases in the intervention district, compared to the control area. Xpert Ultra *trace* positive results accounted for a high proportion of laboratory confirmations in the ACF cohort (51.1% vs 13.7% of those passively diagnosed). Number needed to screen (NNS) to find a TB case differed widely among HCs (NNS:55) and CCs (153). The intervention resulted in an overall increase in TB diagnoses and higher proportion of laboratory confirmation.

## Introduction

Tuberculosis (TB) remains one of the leading causes of death at global scale. In 2020, it was responsible for 10 million episodes of disease and 1.5 million deaths(1). Prompt diagnosis is critical for TB control, as it enables early treatment which in turn, reduces transmission and improves treatment outcomes(2). However, underdiagnosis and underreporting are common in TB surveillance systems (3) and high detection gaps are reported in various WHO regions (2). In 2020, it has been estimated that over 4 out of 10 TB cases were not diagnosed and/or reported to the health authorities(1).

TB diagnosis under National TB Programs (NTP) routine activities in high-burden countries relies predominantly on passive case-finding strategies (PCF). They require individuals to recognize their symptoms and attend a healthcare center to be diagnosed by the health care workers. Conversely, active case-finding (ACF) allows the detection of asymptomatic cases or symptomatic non-healthcare-seeker cases through systematic screening and evaluation of populations at high risk of developing TB disease(4). ACF strategies constitute a useful attempt to reduce TB detection gaps. Unlike PCF, early TB detection in these populations can halt TB transmission, improve disease outcomes due to diagnosis in the early stages of the disease and modify risk perceptions and healthcare-seeking behaviors(5). However, ACF interventions are extremely diverse (6–11) and often tailored to different population characteristics and resources. In fact, ACF effectiveness is inconsistent and their impact on TB control and patient outcomes needs to be rigorously evaluated (12).

Mozambique is one of the high TB, TB-MDR, and TB/HIV burden countries. In 2017, the estimated detection gap was 48% (13). Although the TB detection rate may have increased in recent years, coinciding with the new burden estimates from a national TB prevalence, there are some concerns on the quality or TB diagnostic and reporting services, given the worrying low bacteriological confirmation rate among notified cases(2). Several factors could explain a low case detection, including health system structural limitations and health-seeking behavior patterns(14), probably derived from low health risk perception (15), traditional health beliefs, and poor diagnostic infrastructure (14,16). Besides, diagnosis in people living with HIV (PLHIV) is challenging (17). This might have greatly contributed to the low case detection rate (18). Thus, the implementation of ACF interventions in high-risk populations could contribute to reduce the burden of missing cases.

Household members and people who interact frequently with a TB case (household contacts, HCs or community contacts, CCs) are at higher risk of TB than the general population, due to the longer time of exposition(19). As the bacillary load of index cases (IC) could be a proxy indicator of infectiousness (20,21), IC’s bacillary load could be used to stratify transmission risk. In 2017, Xpert MTB/RIF Ultra appeared as more sensitive version of the previous Xpert MTB/RIF. However, by 2018, the national TB programme (NTP) had not scale-up its routine use. Thus, we designed an innovative ACF strategy, based on the combination of Xpert Ultra semi-quantitative results and spatial parameters on tuberculosis case notifications (microbiologically confirmed and overall) in a high TB and HIV burden district of Southern Mozambique. The main objective of this analysis is to assess the impact of this combined intervention on TB case detection.

## Methods

### Study design, population and setting

This is a prospective controlled intervention study, which evaluates the effects of an ACF strategy on the number of TB cases notified in the district of Manhiça (intervention area) over a period of 12 months (January-December 2018). Manhiça is a semi-rural area, 80 kilometers away from the capital, with a population of approximately 204,953 inhabitants as of 2019(20). The study was implemented by the Manhiça Health Research Center (CISM, from its acronym in Portuguese), which runs a Health and Demographic Surveillance System (HDSS) since 1996. This HDSS routinely collects sociodemographic information and actively follows important demographic events of nearly 100% of the population in the district (20,21).

A control area was used in order to assess whether the trends observed in the district of Manhiça could not be explained by natural variations of the disease in the absence of intervention. Notification data of 2 rural districts in Maputo province, that do not limit with our district were used (Namaacha and Matutuine). Those districts did not participate in other ACF projects in place or implemented the use of Xpert MTB/RIF or Xpert Ultra during 2018 or in previous years.

### Xpatial strategy

During the study period, the novel Xpert MTB/RIF Ultra assay (Cepheid, Sunnyvale, CA, USA, hereinafter Xpert Ultra) was used as the frontline test for active TB diagnosis in the entire district, as it has been described elsewhere (22). New and relapse TB patients who started treatment from January 1st to December 31st of 2018 in Manhiça district were invited to participate in this study. Those providing informed consent were included as index cases (IC) of the strategy and followed the study algorithm shown in Figure 1.

**Figure 1.**
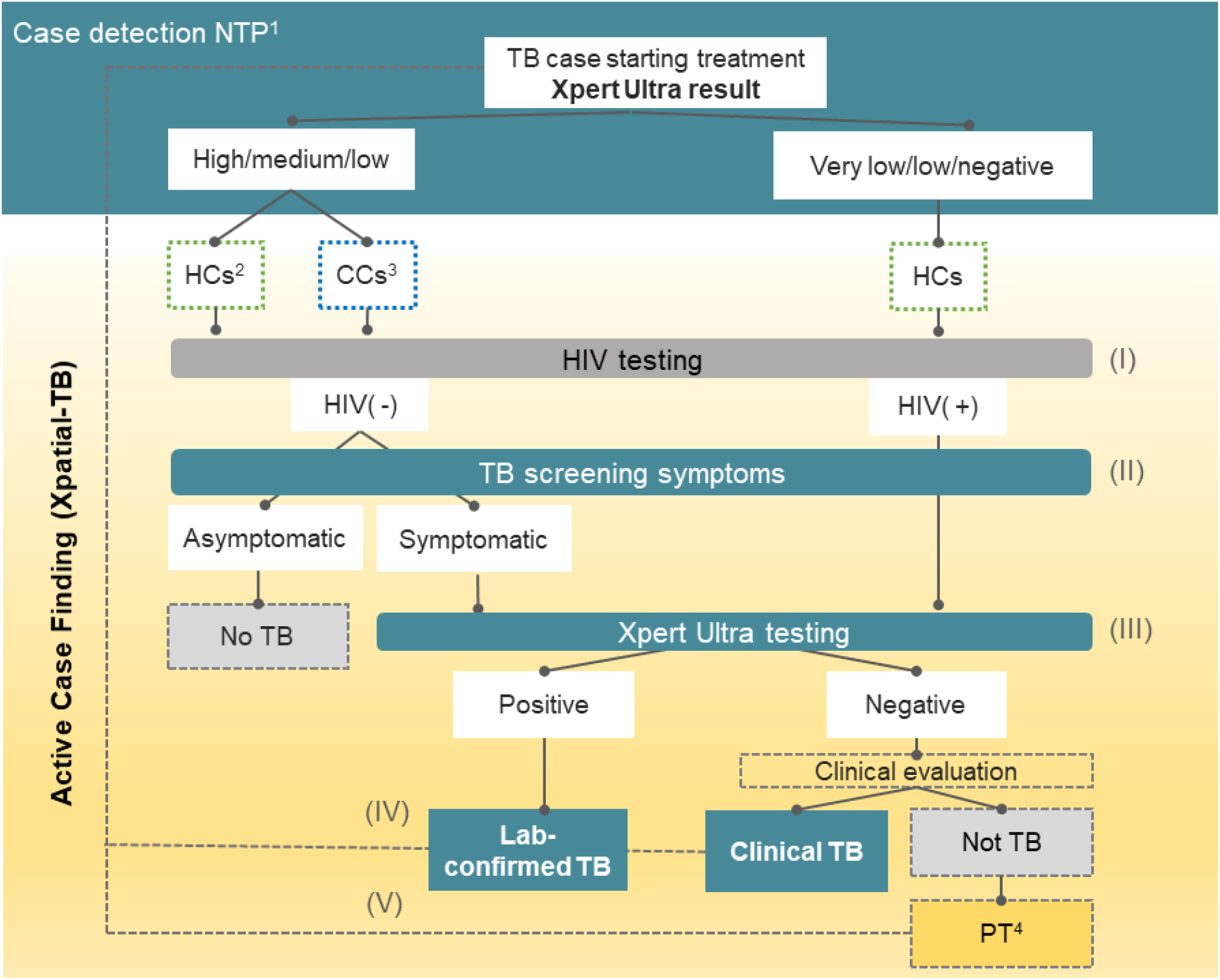
Xpatial algorithm. ^1^ NTP: National TB Programme; ^2^HCs: household contacts; ^3^CCs: community-close contacts; ^4^PT: Preventive therapy The Active Case Finding Intervention had 5 steps: I) HIV testing; (II) Screening of TB symptoms; (III) Ultra testing for those accomplishing criteria (HIV positive regardless symptoms or symptomatic contacts); (IV) identified TB cases were referred to the NTP to started on treatment; (V) children < 5 years old were referred to started preventive therapy.

#### ACF activities

Once and IC was identified, a list of household and neighboring contacts (eligible contact population) was generated based on HDSS registers.

The contact population identified per IC depended on two main variables: the bacillary burden of the IC, and the household density of the neighborhood where the IC lived in. The bacillary burden was classified as per Xpert Ultra semiquantitative results (High/Medium/Low/Very low/Trace/Negative). Following our predefined algorithm, ICs with low, medium or high Xpert Ultra results, generated both household (HCs) and community contacts (CCs). Those ICs with a very low, trace or negative result, only generated HCs (based on studies relating infectiveness and bacillary burden 21,22). The area around which neighboring contacts were screened depended on predefined radii from the ICs’ household. These radii were pre-established based on the household density of the neighborhood where the IC lived in.

After IC enrolment, three field teams composed of one nurse and one field worker each, visited the contacts identified though the HDSS. Up to three attempts (visits) were made within 90 days of IC enrolment in order to recruit identified contacts. After providing informed consent, contacts were systematically screened following a 3-step approach: 1) HIV status verification: All participants with unknown or negative HIV status were offered HIV testing; 2) TB symptom screening (22) (cough, hemoptysis, fever, weight loss and night sweats); and 3) sputum collection for microbiological testing for HIV negative participants who reported TB symptoms and HIV positive participants irrespective of symptoms (Figure 1). If participants were not able to provide spontaneous sputum, field sputum inductions were performed using portable nebulizers. Symptomatic patients were followed at the study facilities located in the Manhiça District Hospital. If clinical or lab-confirmed TB diagnosis was made, participants were referred to NTP facilities for treatment initiation. In case of pediatric contacts less than 12 years of age, additional TB symptoms included failure to thrive or reduced playfulness. If TB symptoms were present, attempts were made to obtain induced sputum, gastric aspirates, or, if available at the time of visit, urine of stool samples, who were later processed for Xpert Ultra.

A more detailed description of the contact ratio calculations, diagnostic samples collection and laboratory methods can be found in the supplementary material (S1).

#### Data analysis

Statistical analysis was performed using R version 3.5.2 (R Foundation for Statistical Computing, Vienna, Austria). The effect of the intervention was evaluated by: i) assessing the TB care cascade and describing process indicators; ii) calculating numbers needed to screen (NNS) to find one case of TB as indicator of undetected TB and efforts required to diagnose one case among different groups and epidemiological situations (NNS = 1/prevalence(23)); iii) conducting an interrupted time series analysis with and without control (CITS/ITS), of aggregated quarterly TB notification cases.

### *ITS/CITS models* (24)

Two periods were set: the pre-intervention (2015-2017) and the intervention period (2018). The pre-intervention period was used as a first counterfactual, to make 2018 predictions in the absence of the intervention (before and after comparison). In addition, the control area was used as a second counterfactual. We assumed no seasonality in TB notifications. The outcomes of interest were: number and notification rates of TB and laboratory-confirmed TB cases. Population counts for Manhiça district were obtained from the HDSS, and was extracted from national projections census data for the control area.

We employed segmented linear regression models. The following equation was applied for the complete model:

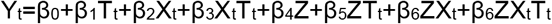

Where Y_t_ is the outcome at given point, T is time variable, X is a dummy variable representing the intervention (preintervention period as 0; intervention period as 1); and Z represents the intervention cohort (control area=0; intervention area=1). Model coefficients (β) were used to estimate the impact of the intervention by measuring outcome levels, trends and step/slope changes depending. Further details can be found in Supplementary material (S2). Autocorrelation was tested by Cumby–Huizinga general test. Estimates were reported with standard errors and p-values as measures of precision. The final model was used to estimate a counterfactual of the notified cases in Manhiça district if the intervention had not occurred and difference with final cases reported.

Additionally, we assessed differences in the distribution of TB cases (detected by PCF or through our ACF strategy in both HC and CC) by age, sex, sociodemographic distribution, HIV status, bacteriological confirmation, Xpert Ultra results, TB type and X ray results. Continuum variables were compared through means t-test and categorical variables through chi-square test.

#### Ethics considerations

The study was approved by the National Bioethics Committee for Health of Mozambique (CNBS, Ref:369/CNBS/17) and the Internal Bioethics Committee of CISM. All methods were performed in accordance with the relevant guidelines and regulations. All study participants (IC, HC and CC) signed written informed consent after a verbal explanation and written information about the study was provided. For participants under 18 years of age, an informed consent was obtained from their relatives, parents or guardians.

## Results

### Process indicators (Figure 2)

From January to December 2018, a total of 1010 new or relapse index TB cases, starting treatment, were included in the study. Following the Xpatial algorithm (Figure 1), an eligible population of 4,394 HCs and 6,373 CCs was identified by the HDSS; of those, 72.0% (3165/4394) and 74.2% (4730/6373) were respectively verbally screened for TB symptoms. A total of 17.0% (1345/7895) were eligible for sampling (see Methods): 10.7% of all screened contacts (852/7812) who presented TB related symptoms (15.8% HCs vs 7.5% CCs, p< 0.001) and PLHIV without symptoms, which accounted for 6.2% (493/7812) of all contacts screened. We obtained sputum from 52.9% of them (712/1,345). Around 1.1% of all contacts screened (89/7,895) refused HIV testing.

**Figure 2.**
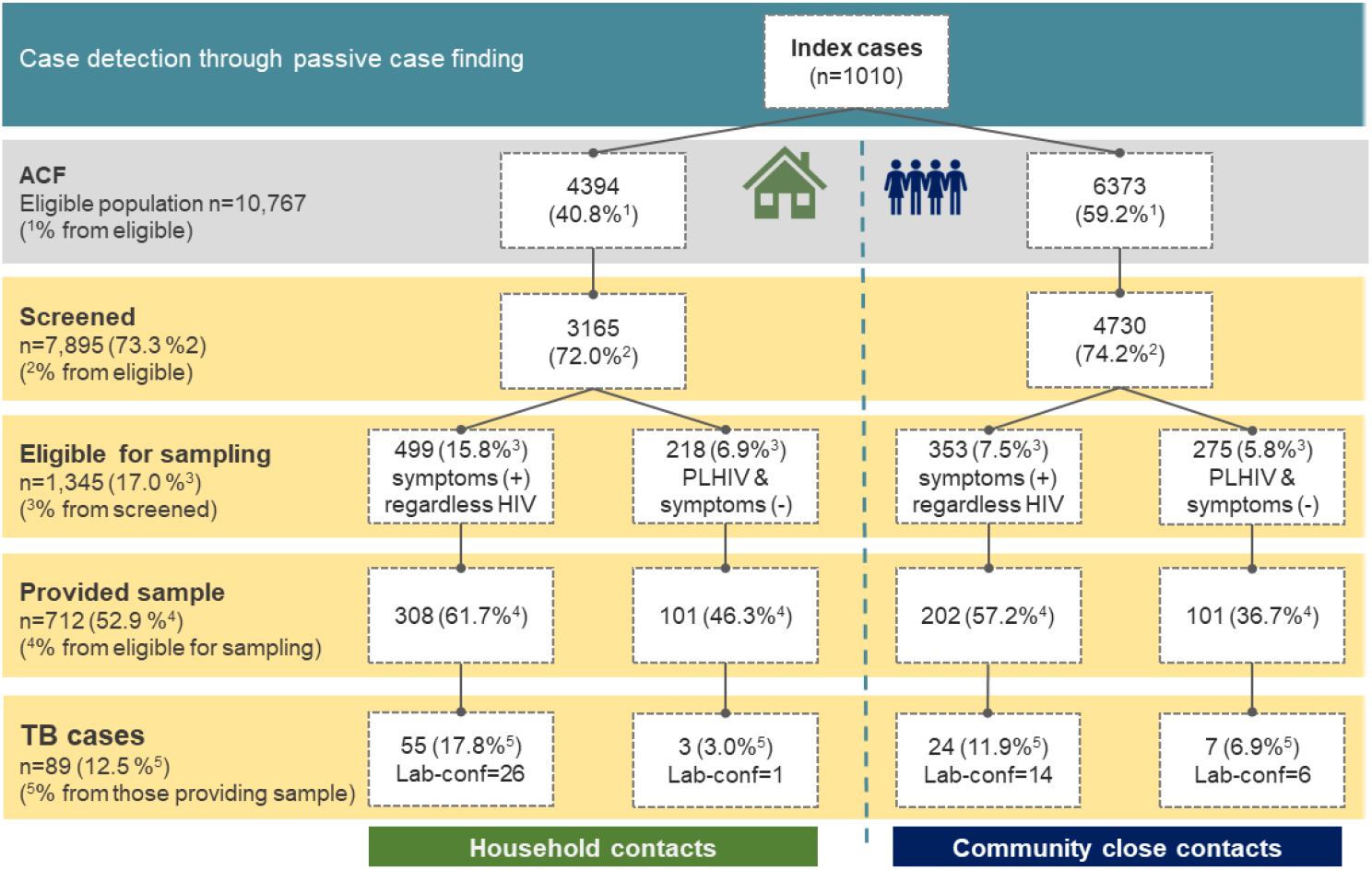
Process indicators; PCF: Passive Case Finding; ACF: Active Case Finding; Microb conf: microbiologically confirmed.

As a result of the ACF strategy, 89 contacts were diagnosed with pulmonary TB, most of them symptomatic (79/89, 88.8%), but also 10 (11.2%) asymptomatic PLHIV. All of them started treatment. There was an overall microbiological confirmation of 7.8% (40/510) among symptomatic contacts who provided sample, and 3.4% (7/202) among asymptomatic PLHIV.

The overall NNS to find a TB case was 88.7 cases, 54.6 for HCs and 152.6 for CCs. This indicator widely varied among specific groups (Table 1). It was necessary to screen 168 individuals to find 1 lab-confirmed case, 99.9 to find a symptomatic case, and 190.8 to find a paediatric case. Overall, NNSs were lower for HCs, except for asymptomatic PLHIV. Absolute numbers to calculate NNS are displayed in Supplementary material (S3. Table 1).

**Table 1.** Number needed to screen for different study populations

|  | HCs<sup>1</sup> | CCs<sup>2</sup> | total |
| --- | --- | --- | --- |
| NNS<sup>3</sup> overall | 54.5 | 152.6 | 88.7 |
| NNS<sup>4</sup> lab-confirmed | 117.2 | 236.5 | 168.0 |
| NNS<sup>5</sup> symptomatic | 57.5 | 197.1 | 99.9 |
| NNS<sup>6</sup> asymptomatic PLHIV | 1055.0 | 675.7 | 789.5 |
| NNS<sup>7</sup> paediatric | 89.6 | 1102.0 | 190.8 |
| NNS<sup>8</sup> women | 113.0 | 236.5 | 164.5 |
| NNS<sup>9</sup> men | 105.5 | 430.0 | 192.6 |
| NNS<sup>10</sup> women among women | 54.5 | 96.9 | 72.2 |
| NNS<sup>11</sup> men among men | 54.7 | 253.7 | 108.1 |

Footnote: 1) HCs: Household contacts; 2) CCs: community close contacts; 3) NNS: number needed to screen to find a TB cases in the overall study population; 4) number needed to screen to find a laboratory—confirmed cases; 5) NNS to find a symptomatic patient; 6) number needed to screen to find an asymptomatic coinfecting TB/HIV patient; 7) NNS to find a patient < 12 years old ; 8 ) NNS to find a women with TB; 9) NNS to find a men with TB ; 10) women needed to screen to find a TB case; 11) men needed to screen to find a TB case

### Characteristics of cohorts

#### Sociodemographic characteristics of index cases (ICs) (Table 2)

In 2018, 1020 incident TB cases were reported by the NTP. Individual information of those cases identified passively was available in 1010 cases (signed informed consent); 53.7% (543/1010) were male, the median age was 39.5 [IQR 30.1-52.4], being the age distribution slightly different for women and men. PLHIV accounted for 63.5% of ICs (641/1010), and 92.7% (936/1010) presented pulmonary disease. Overall, 37.2% of cases (376/1010) were microbiologically confirmed, 62.1% (233/376) of them being PLHIV. This translated into 36.4% (233/641) of bacteriological confirmation within this specific population, similar to the proportion of HIV-negative confirmed patients (38.6%, 142/368). Of those bacteriologically confirmed by Ultra, 58.5% (209/357) fell into the high or medium categories and 13.7% (49/357) into the ‘trace’-call. Paediatric cases (children under 12 y.o) accounted for 6.1%. At the end of the study period, 10.1% (102/1010) of TB cases died. Just 1.1% (11/1010) showed resistance to rifampicin by Ultra testing. Further details on ICs characteristics are shown in Table 1

**Table 2.**
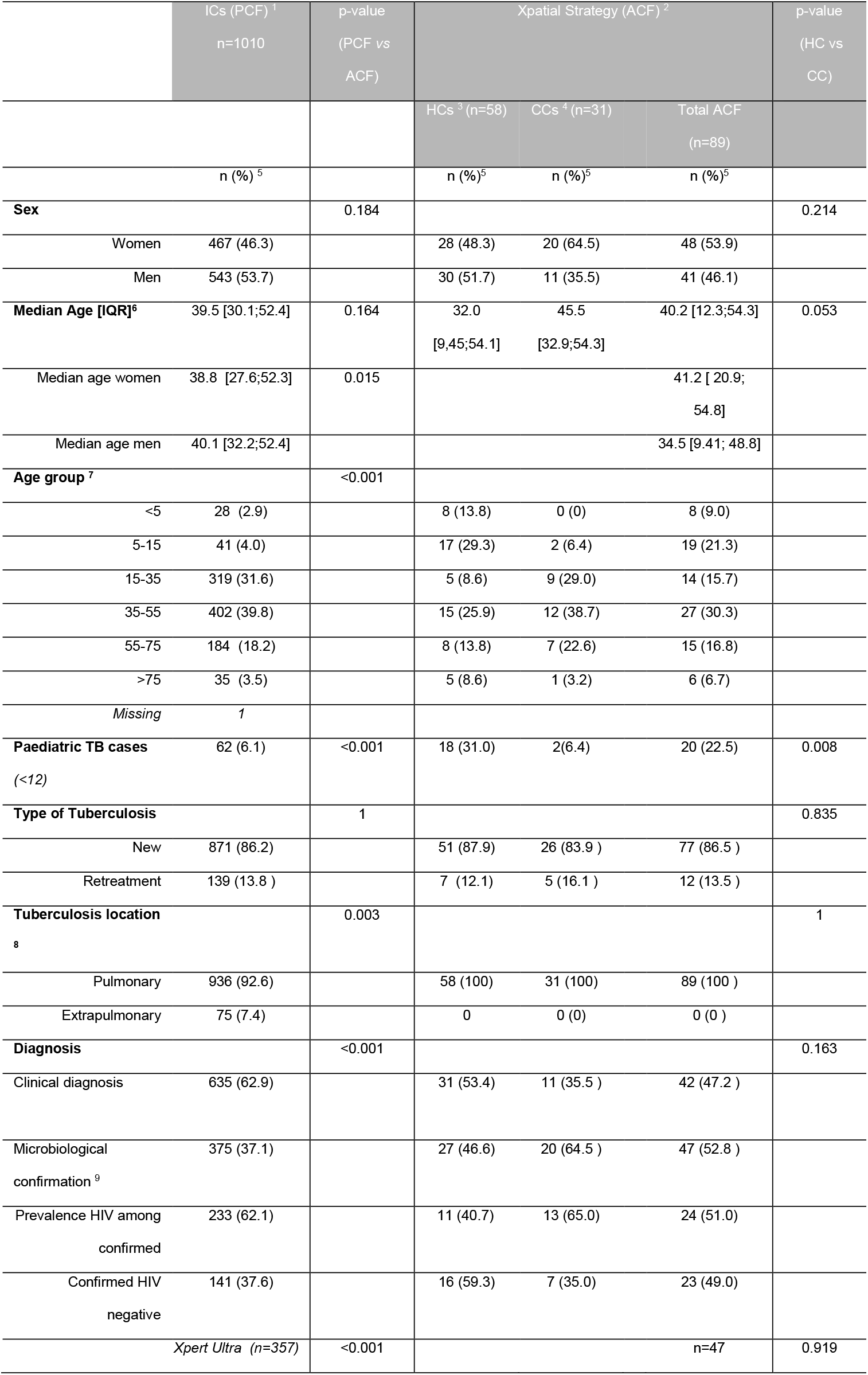
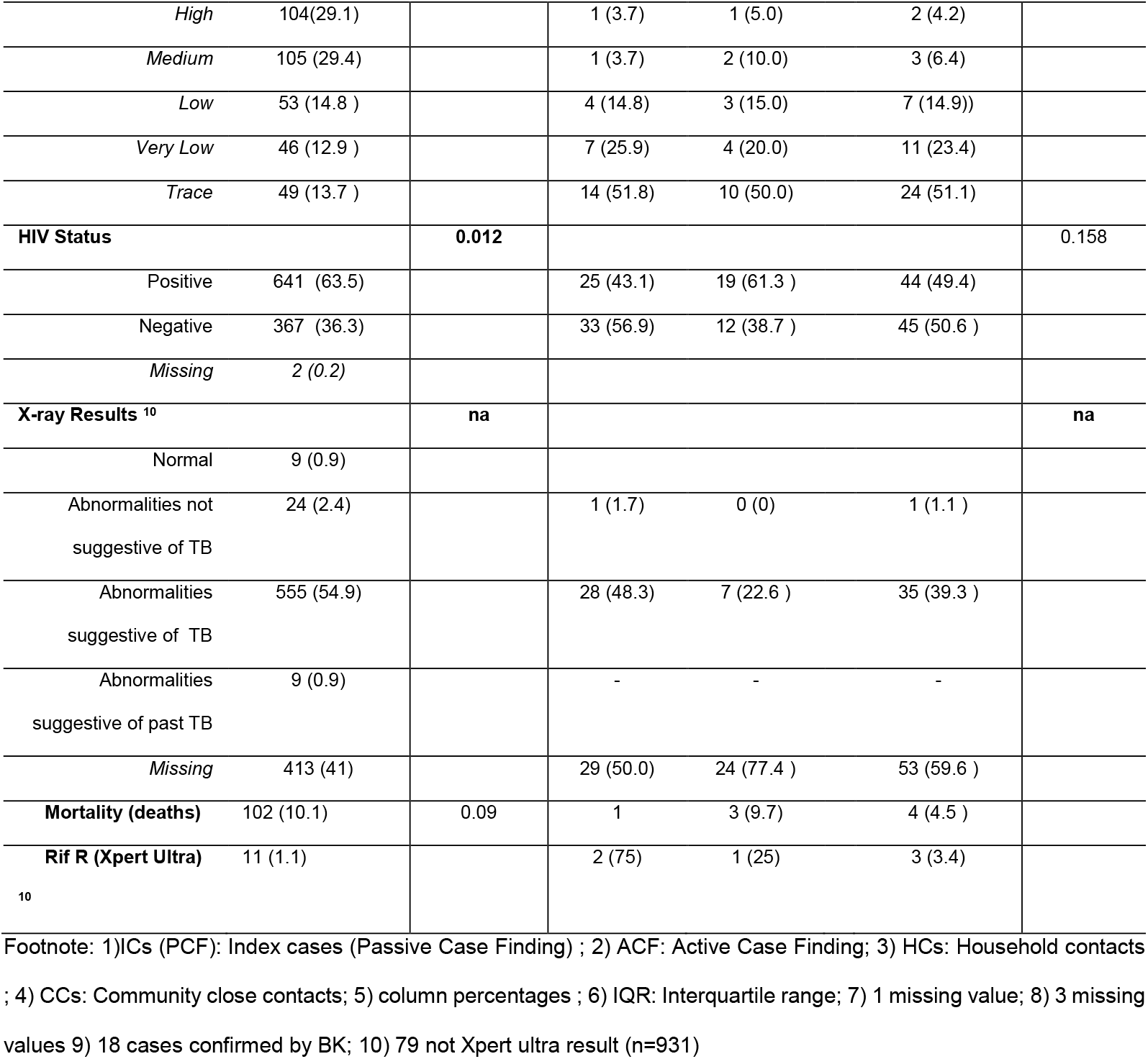
Characteristics of index cases and TB cases diagnosed through the Xpatial strategy (overall and stratified by type of contact).

#### Sociodemographic characteristics of screened population (Supplementary Table 3)

From those 1010, 7895 people were screened, and 55.7% (4398/7895) of them were women. The median age was 15.5 [8.0;32.4] being the age distribution different between HCs and CCs. PLHIV accounted for 8.9% (701/7895) of evaluated contacts. This percentage was higher in HCs than in CCs (10.9% vs 7.5% p<0.001). More than one third of screened contacts (35.1%) belonged to Manhiça village health area.

#### Sociodemographic characteristics of TB cases identified by the Xpatial intervention (Table 1)

From the total of 89 TB cases identified, 53.9% (48/89) were women. The median age was 40.2 [IQR:12.3; 54.3]. The age of cases diagnosed from HCs was significantly different from CCs (32.0 IQR [9.4;54.1] vs 45.5 [32.9;54.3], p-value=0.05). Pediatric cases were found predominantly in household contacts (31.0% HCs vs 6.4% CCs); 25/27 children <15 and all children < 5 years old were found in household screening. The prevalence of HIV coinfection in this cohort was 49.4% (44/89), and 51.0 % (24/47) among confirmed patients, being higher for HCs than CCs (59.3% vs 35.0%). Most TB cases had no previous history of TB (77/89, 86.5%).

More than half of cases was bacteriologically confirmed (52.8%, 47/89), and 74.5% (35/47) of them relied on Ultra’s lower categories (very low or trace). Three cases showed resistance to rifampicin (3.4%) and 4 patients died (4.5%) during follow-up.

Thirteen cases identified through PCF (9 women, 4 men) shared household with previously diagnosed PCF cases but were not detected by the ACF strategy. From those, only 9 had been registered as contacts; 2 were not reached, 4 were HCs screened and 1 CCs.

### Notification impact

As six ACF cases were diagnosed between January and March 2019 (window period), just 83 cases were used for the rate trend analysis.

#### ITS

Evaluation of the implementation of Xpert Ultra as the frontline test in the district of Manhiça

As shown in Figure 3, between 2015-2017 (pre-intervention period) notification rates (NR) in Manhiça district showed a slight growing pattern up to 2017 (blue lines). Notifications then declined throughout 2017. This trend was reverted in 2018 (intervention period). Conversely, the proportion of laboratory confirmed cases was consistently low before the intervention (average of 23.5% in 2015-2017), showing a declining trend that was sharply reversed during the intervention (37.2%(375/1020)).

**Figure 3.**
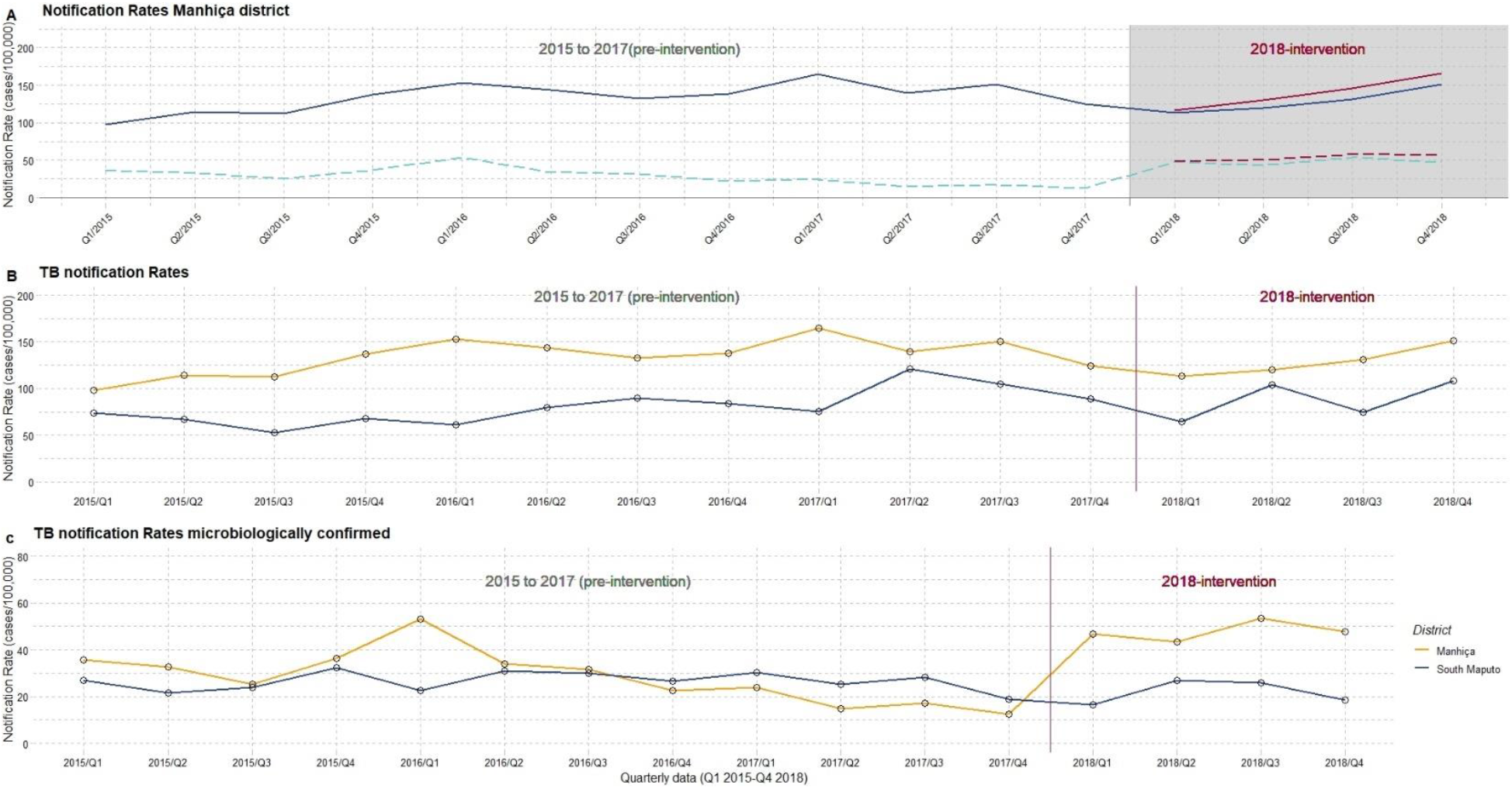
**A)** TB notification rates in Manhiça district pre-intervention/during intervention. TB cases excluding those derived from the ACF intervention are displayed in dark-blue. Additional ACF-cases are displayed in red. Bacteriologically confirmed cases are drawn in light-blue dash lines. **B**) TB Notification rates in Manhiça (blue) and the control area (Namaacha and Matutuine, yellow) **C**) Notification rates of microbiologically confirmed cases in Manhiça and the control area

The ITS (interrupted temporal analysis for Manhiça district, Table 3, Supplementary S5 and S6), estimated a pre-intervention positive trend among notified cases, with a small increase in quarterly NR (β_1 =_3.09 cases/100,000, SE=1.24, p-value=0.003). Although at the beginning of the intervention there was a reduction in the number of reported cases (-168/100,000, p-value=0.1, following the downward trend which started in 2017, Figure 3), it was thereafter followed by an increasing NR (β_3_ difference pre-post intervention=9.54 /100,000)(Table 3).

**Table 3.**
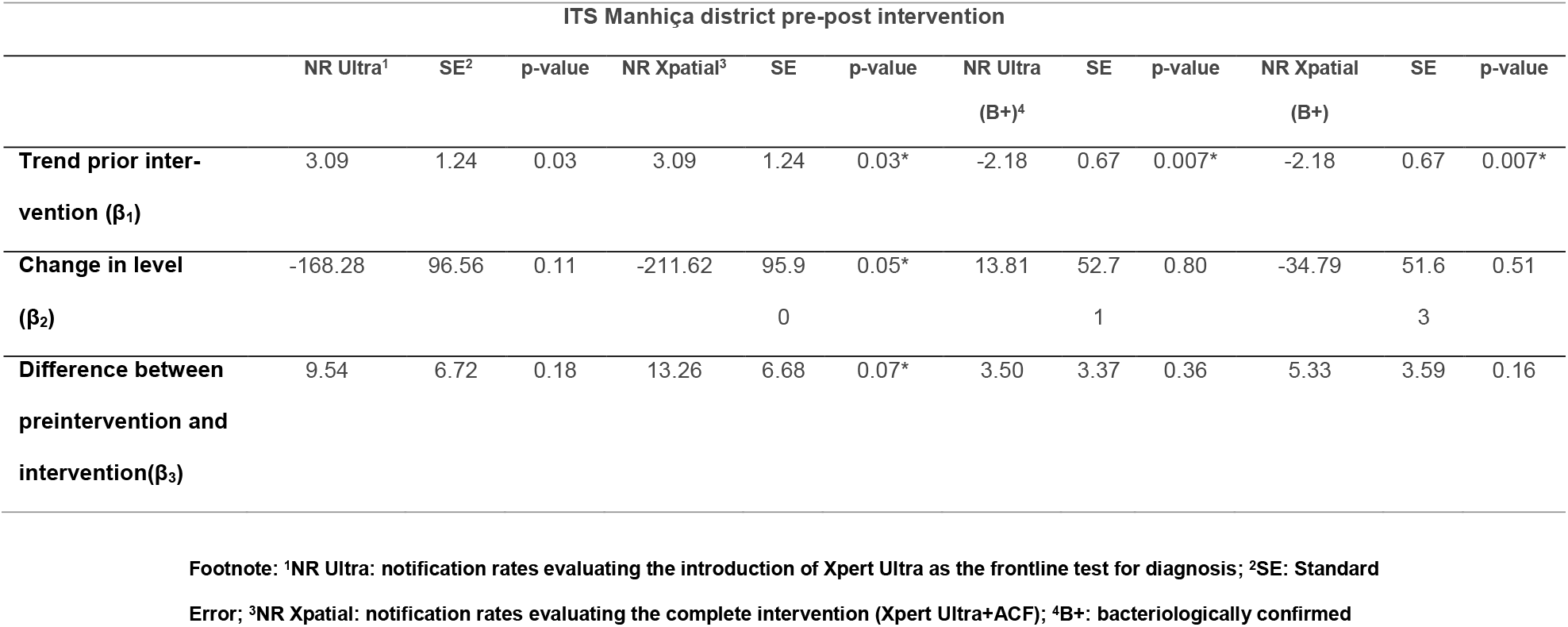
Comparative of ITS model parameters for quarterly case notification rates for all cases and jus bacteriologically confirmed TB cases, evaluating both, the implementation of Xpert Ultra as a frontline test for PCF and the combination with ACF activities.

The ITS confirmed a marked pre-intervention decline in the number of bacteriologically diagnosed cases (Supplementary S6); (β_1 =_-2.18/100,000 per quarter, p-value=0.007). This trend shifted during the intervention, with a positive difference (β_3_) of 3.5 lab-confirmed cases /100,000 per quarter (p-value=0.36)

#### ITS

Evaluation of the implementation of the Xpatial intervention (Ultra + ACF)

The ACF intervention further increased by 8.2% all forms of TB cases detected in 2018 (83 ACF/1010 PCF) (Figure 3). The final proportion of bacteriologically confirmed TB cases during the intervention period was 38.3% (422/1103). ITS model parameters estimated a difference of 5.33 cases bacteriologically confirmed/100,000 population between the pre-intervention and the intervention period (Table 3, Supplementary S6 and Figure 4)

**Figure 4.**
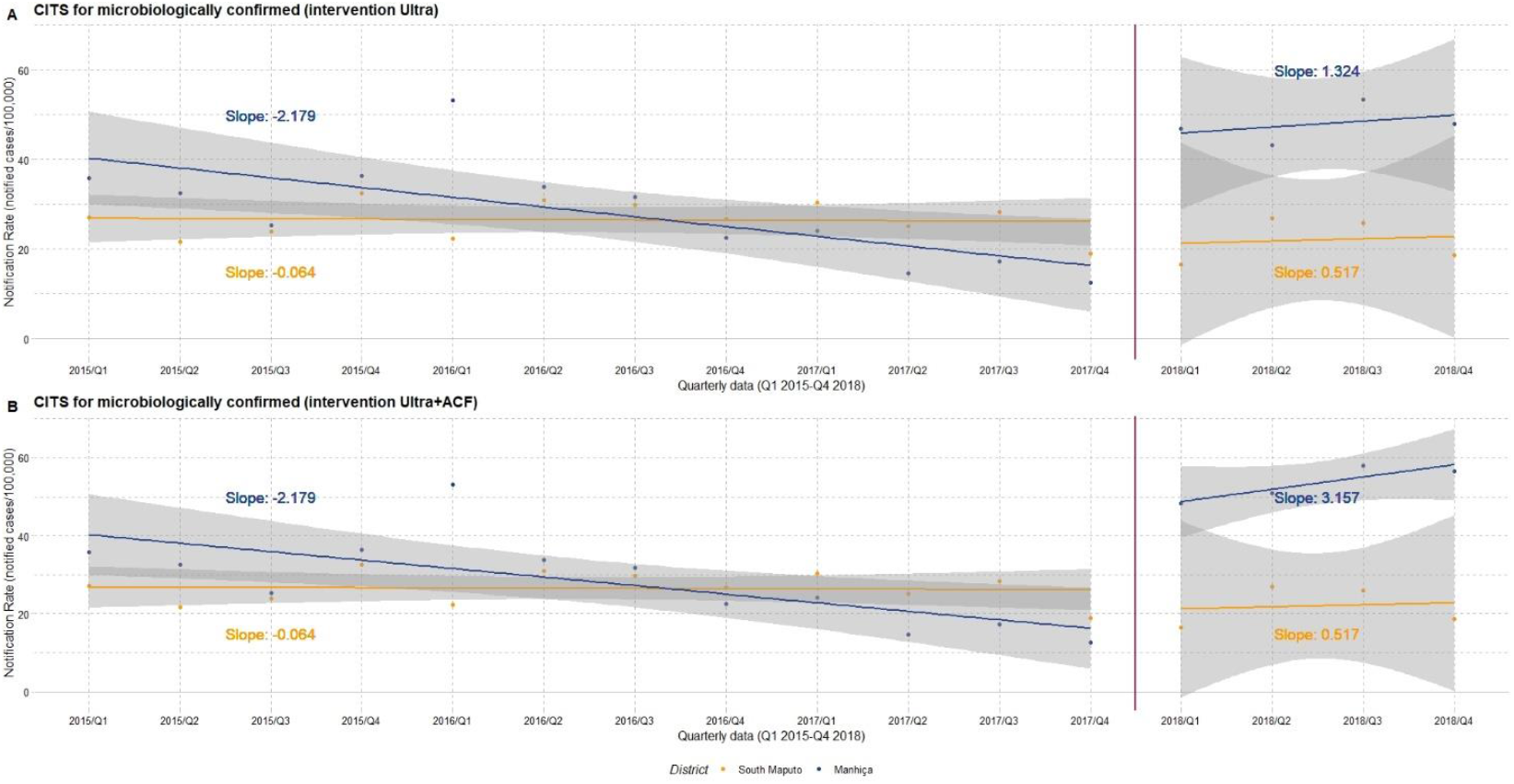
Comparative impact of the intervention using a controlled interrupted series analysis (CITS) (control area, Namaacha and Matutuine, South Maputo).

#### CITS

Evaluation of the implementation of the Xpatial-TB intervention (Ultra + ACF) and comparison with the control area (Table 4 and Figure 4)

**Table 4.** Comparative of CITS model parameters of quarterly notification rates for all cases and just bacteriologically confirmed TB cases, evaluating both, the implementation of Xpert Ultra as a frontline test for PCF and the combination with ACF activities, in comparison with the control area (control Districts)

| CITS comparing Manhiça and the control area pre-post intervention |  |  |  |  |  |  |  |  |  |  |  |  |
| --- | --- | --- | --- | --- | --- | --- | --- | --- | --- | --- | --- | --- |
|  | NR Ultra<sup>1</sup> | SE<sup>2</sup> | p-value | NR Xpatial<sup>3</sup> | SE | p-value | NR Ultra<br>(B+)<sup>4</sup> | SE | p-value | NR Xpatial (B+) | SE | p-value |
| **Difference between preintervention and intervention trend, control ($\beta_3$)** | 6.49 | 6.79 | 0.71 | 6.49 | 6.79 | 0.71 | 0.58 | 3.01 | 0.19 | 0.58 | 3.01 | 0.19 |
| **Difference in baseline intercepts ($\beta_4$)** | 58.46 | 12.99 | <0.001 | 58.46 | 12.99 | <0.001* | 15.58 | 5.76 | 0.012* | 15.58 | 5.76 | 0.012* |
| **Difference in pre-intervention trends ($\beta_5$)** | -0.77 | 1.76 | 0.66 | -0.77 | 1.76 | 0.66 | -2.11 | 0.78 | 0.012* | -2.11 | 0.78 | 0.012* |
| **Difference in intervention step change ($\beta_6$)** | -50.68 | 137.87 | 0.72 | -94.01 | 137.4 | 0.5 | -1.39 | 61.18 | 0.98 | -22.37 | 60.24 | 0.71 |
| **Difference in intervention trends ($\beta_7$)** | 3.06 | 9.6 | 0.75 | 6.77 | 9.57 | 0.49) | 2.92 | 4.26 | 0.50 | 4.75 | 4.19 | 0.27 |

Footnote: <sup>1</sup>NR Ultra: notification rates evaluating the introduction of Xpert Ultra as the frontline test for diagnosis; <sup>2</sup>SE: Standard Error; <sup>3</sup>NR Xpatial: notification rates evaluating the complete intervention (Xpert Ultra+ACF);
<sup>4</sup>B+: bacteriologically confirmed

For the CITS, the regression model identifies no overall significant-level or trend change between the control and the intervention area during the intervention period. The initial down step in number of cases reported occurred in both districts, as well as the recovery, although the decrease in NR was higher in the control area (difference step change *β*_*6*_ of -50.68).

On the contrary, a significant difference in pre-intervention trends for microbiologically NR between areas was confirmed (-2.18 Manhiça vs -0.06 South Maputo, p-value=0.012). This trend was positively reverted during the intervention period in Manhiça district (Figure 4), although the model did not show statistical significance.

## Discussion

To our knowledge, this was one of the largest ACF study conducted in household and community contacts in sub-Saharan Africa using Xpert Ultra as the frontline screening test, and which included both, spatial and microbiological parameters to select a target population. In addition, we assessed the impact of using Xpert Ultra under programmatic conditions. The study shows: a) an overall increase in the number of lab confirmed cases and in the notification rate of microbiologically confirmed TB in the Manhiça district during the study period; b) a higher than expected proportion of paediatric cases diagnosed through the ACF strategy; c) a higher proportion of microbiologically-confirmation among TB cases diagnosed through ACF activities (compared to those diagnosed in PCF); and d) an increased number of Ultra positive results with low semiquantitative Xpert results among ACF cases (compared to PCF cases).

Undoubtedly, the integration of Xpert Ultra on programmatic case detection had a positive impact on the notification rate of laboratory confirmed cases and reversed the pre-intervention downward trend in the Manhiça district. Compared to the pre-intervention period, the proportion of lab-confirmed cases changed from 23.5 to 37.2% (excluding cases arising from ACF). The use of a more sensitive test for TB diagnosis is likely be the main reason for the considerable difference in lab-confirmed cases in 2018. Indeed, 13.7% of notified TB cases reported on Ultra’s new lowest category, “trace”. If the former Xpert assay had been used instead, it is likely that those cases would have not been confirmed (and a proportion of them would have been not diagnosed for TB).The initial debate on the significance of trace results for the Xpert MTB/RIF Ultra cartridge seems to have achieved a certain consensus. The latest recommendations advocate against retesting in favor of a cautious assessment of the clinical presentation and individuals’ characteristics.(25)

The overall proportion of microbiological confirmation in 2018 was higher than in the pre-intervention period. It was also considerably lower in the PCF cohort (37.1%), and aligned with the annual country report (39%), when compared to the ACF cohort (52.8%) (26). The low confirmation rate is been repeatedly questioned by the World Health Organization since the national TB prevalence survey(2). If a widely-applied screening found more lab-confirmed cases, we may suspect a large number of hidden and missed patients in the district. Moreover, the lower bacillary burden found in samples from ACF activities would likely mean that we found patients with less severe case disease, meeting one of the central objectives of screening activities: early detection of cases to interrupt the transmission chain (27)(28).

Although the comparison with the control area did not show significant difference in TB notification (overall or lab-confirmed cases) using CITS analyses, there is a clear shift in the trend and amount of confirmed cases for Manhiça district that did not happen in the control area. It is likely that the few data points available from the NTP reports (quarterly data, four points), might have played a role in the absence of statistically significant results. Similar to other ACF intervention studies, we used a linear regression to model the data. However, we could not include other covariates that might have affected the diagnosis of cases and explain irregular reporting over time (stock-outs of reagents, health coverage or retention in care data). A higher number of control districts, and the assessment of post intervention trends, would have helped to formally elucidate whether our intervention had a positive impact in CITS analyses.

Prior to the study, ACF activities were inconsistent in the district and relied on the availability of community-health workers. The implementation of a massive screening strategy, targeting pre-defined high-risk populations increased the overall number of detected cases by 8.9%, and the number of laboratory-confirmed cases by 15.7%. Evidence on the effectiveness of community-based ACF activities vary widely among studies (29), and determining whether our strategy was affordable and cost-effective would depend on economic evaluations (23). Nevertheless, process indicators and differences found among screened/PCF/ACF cohorts may provide information on likely sources of detection gaps in our district and populations who would benefit the most from the intervention.

The effectiveness of HCs screenings was considerably higher than for CCs. NNSs were higher for CCs than for HCs in almost all cases (i.e overall NNS to find a TB case was 54.6 HCs vs 152.6 CCs). Those differences were even higher among paediatric cases (NNS HCs 89.9 in HCs vs NNS 1102 in CCs). Since TB disease among children results from persistent close contact with an infectious IC, this finding would support the benefit of intensive household screening and confirm the low paediatric TB case detection in our setting, as previously described(30). Reported data on childhood TB is an important indicator when evaluating NTPs. For the same year, Mozambique reported that 13% of cases were under 15 years old, far from the 6% identified in the district. This reinforces the need to take urgent actions to increase TB detection in this vulnerable population, such as integrated TB care interventions of the NTP and maternal and child health programmes(31)(32).

The prevalence of HIV/TB cases identified through the NTP was over 60%, matching previous reports (17). These figures are consistently higher than those reported for the country as a whole (36%) and justified (at the time when the study was designed) the strategy of universal testing for HIV individuals regardless of symptoms. Surprisingly, the number of TB/HIV cases found was lower than expected for this specific population (overall NNS of 789.5) and far from that reported in similar studies (33). Likewise, the prevalence of HIV among screened contacts was also lower than previously seen, and specific analysis are being conducted to better understand the HIV burden in several populations of the district. Of note, the specific NNS for finding females and males was similar among HCs, but differed significantly in CCs (NNS to find a man: 430 *vs* NNS: 226 to find a woman). Although the local population pyramid (ratio F/M 1.2) (34), would skew the sex distribution of screened individuals and ACF cases (54% of females), this did not happened for HCs. Therefore, further gendered research should be conducted to gain better understanding on the role of CCs screening in finding more women.

Given the high NNS for CCs and the low yield reached on community paediatric cases, we wondered whether TB cases identified through CCs were actually cases epidemiologically linked to the IC (or, alternatively, co-prevalent cases). Although the district has some populated areas, most inhabitants live in rural, open and spacious compounds. Prolonged daily interaction with infected people may take place in other places, such as public transport, bars or workplaces(35). In addition, although the study might have played a key role in Manhiça’s uptake and integration of the new Ultra testing to routine care, we lack consistent data to evaluate post-intervention years. Furthermore, the number of cases identified through the ACF strategy was lower than initially expected. It can be argued that the small window screening period used in this intervention (3months), might be too short to find recent contacts or older primary cases(35). Molecular epidemiology studies should shed more light on the nature of the link between IC and TB cases found among contacts (36)(37)(38). Other important questions such as what the best window period for screening is, whether there is a need for time-separated screening visits, the role of HIV status in TB transmission and what role gender plays on the impact of ACF activities need to be answered to refine ACF intervention designs. Lastly, TB control might no longer be based on the historical dichotomy, either active or metabolically inactive latent infection (39). If subclinical cases can transmit the infection in the community, traditional contact-tracing interventions based on TB compatible symptoms-should be expanded to testing asymptomatic, HIV negative contacts.

### Conclusion

Through the implementation of a combined strategy, the scaling-up of the Ultra test for programmatic activities and the implementation of an innovative ACF strategy, more bacteriologically confirmed TB cases were reported in the district of Manhiça. However, our CITS analysis could not confirm the impact of the strategy on either TB case notification or bacteriologically confirmed cases, potentially due to limitations in the analytic approach and the limited data on confounding factors. Paediatric population benefited the most from the ACF strategy and HCs screening was confirmed as an effective strategy to find microbiological confirmed cases in early stages of the disease.

## Data Availability

All data produced in the present study are available upon reasonable request to the authors

